# Stress in autism (STREAM): A study protocol on the role of circadian activity, sleep quality and sensory reactivity

**DOI:** 10.1101/2023.02.22.23286298

**Authors:** Clara C. Gernert, Christine M. Falter-Wagner, Valdas Noreika, Barbara Jachs, Nazia Jassim, Kathryn Gibbs, Joaquim Streicher, Hannah Betts, Tristan A. Bekinschtein

**Affiliations:** Department of Psychiatry and Psychotherapy, Medical Faculty, LMU Munich, Munich, Germany; Department of Psychology, School of Biological and Behavioural Sciences, Queen Mary University of London, London, United Kingdom; Consciousness and Cognition Lab, Department of Psychology, University of Cambridge, Cambridge, United Kingdom; Autism Research Centre, Department of Psychiatry, University of Cambridge, Cambridge, United Kingdom; Prediction and Learning Lab, Department of Psychology, University of Cambridge, Cambridge, United Kingdom; Independent researcher

## Abstract

Mental health issues are markedly increased in individuals with autism, making it the number one research priority by stakeholders. There is a crucial need to use personalized approaches to understand the underpinnings of mental illness in autism and consequently, to address individual needs. Based on the risk factors identified in typical mental research, we propose the following themes central to mental health issues in autism: sleep difficulties and stress. Indeed, the prevalence of manifold circadian disruptions and sleep difficulties in autism, alongside stress related to sensory overload, forms an integral part of autistic symptomatology.

This proof-of-concept study protocol outlines an innovative, individualised approach towards investigating the interrelationships between stress indices, sleep and circadian activation patterns, and sensory sensitivity in autism. Embracing an individualized methodology, we aim to collect 14 days of data per participant from 20 individuals with autism diagnoses and 20 without. Participants’ sleep will be monitored using wearable EEG headbands and a sleep diary. Diurnal tracking of heart rate and electrodermal activity through wearables will serve as proxies of stress. Those objective data will be synchronized with subjective experience traces collected throughout the day using the Temporal Experience Tracing (TET) method. TET facilitates the quantification of relevant aspects of individual experience states, such as stress or sensory sensitivities, by providing a continuous multidimensional description of subjective experiences. Capturing the dynamics of subjective experiences phase-locked to neural and physiological proxies both between and within individuals, this approach has the potential to contribute to our understanding of critical issues in autism, including sleep problems, sensory reactivity and stress. The planned strives to provide a pathway towards developing a more nuanced and individualized approach to addressing mental health in autism.

## Introduction

Due to the high prevalence of anxiety, depression, and suicidality among autistic individuals, mental health is a pressing theme and one of the top priorities in the field of autism research (1–2). Stress plays a crucial role in mental health, aligning with the prevalent stress-vulnerability model of mental health problems. Adults with autism spectrum disorder (ASD) face substantial challenges in daily living tasks. Numerous studies indicate self-reported heightened stress levels in individuals with ASD compared to non-autistic individuals (3–5). Research suggests a reciprocal relationship between the severity of autism characteristics and perceived stress (3,6), potentially impeding individuals from seeking help or social support when needed. Elevated levels of perceived stress in ASD are linked to poor social functioning, social outcome, and quality of life (3,4,7). Individuals with ASD encounter increased stress across diverse domains, including sensory challenges, which correspond to anxiety levels (8,9).

Indeed, a major source of stress in autism are sensory-processing difficulties (10–12). Approximately 90% of children diagnosed with ASD are estimated to experience atypical sensory perceptions (13). These sensory processing issues persist across the lifespan of individuals with ASD, with a comparable prevalence rate among autistic adults (14–15). Discomfort arising from specific sensory stimuli can trigger self-injurious or aggressive behaviour in individuals unable to articulate their distress. Alternatively, it may lead to avoidance behaviour, demanding substantial effort and time, consequently impacting everyday quality of life. Recognizing the significance of this trait, the latest version of the DSM (16) acknowledges hyper- or hyposensitivity to sensory stimuli in the description of ASD.

Research has uncovered a connection between physiological stress and sensory sensitivity in ASD (17–18). However, the causal mechanism linking sensory over-responsivity, stress, and the severity of autistic and anxiety traits remains unclear (19, 10). Yet, we can draw on findings from beyond ASD showing that sensory processing patterns do also affect sleep quality (20) and evidence showing that sleep disturbances in general are bidirectionally linked to increased levels of anxiety and depression (21). Indeed, sleep difficulties are known to be a very common problem in autism (e.g., 22-26) with 85.6 % of autistic individuals reporting at least one sleep-related concern (25). Furthermore, a positive relationship has been shown between sleep issues and ASD symptom severity (27). In particular, recent large meta-analytic findings showed decreased sleep efficiency and a higher proportion of light-sleep in people with ASD across the lifespan (28–30). Importantly, poor sleep has been shown to correlate with psychological issues, adaptive functioning, and physical health in people with ASD (31). Likewise, individuals with ASD report circadian abnormalities (32).

The findings summarised above converge to the hypothesis of a vicious circle between disturbed sleep and circadian irregularities increasing sensory reactivity increasing stress levels, which arguably can aggravate sleep problems (33). Stress in the life of an autistic person is likely to be complex and specific to the individual, warranting a scrutiny of first-hand accounts. Although the value of first-hand experience accounts by individuals with ASD was proclaimed already several decades ago (34), research beyond case studies or qualitative research is limited. To date, the field of autism research is missing the assessment of nuanced, subjective experiences that can be quantitatively linked to neurocognitive mechanisms. Subjective impressions have predominantly been assessed using Likert scales, questionnaires, or qualitative approaches through interviews. However, Likert scales provide a rudimentary tool to get information about a person’s experience, qualitative approaches are time-consuming, require expertise, and, in practice, the outcome is difficult to compare across participants and even less easy to link to neurocognitive mechanisms.

To overcome these limitations, the current study adopts a quantitative approach to capture complex individual, conscious experiences by applying the Temporal Experience Tracing (TET) method (35). TET allows participants to graphically indicate changes of intensity of different predefined dimensions of experiences. Rather than using Likert scales to rate experiences across categories, individuals retrospectively draw the intensity of their experiences using a grid. The vertical axis represents the intensity of different phenomenal dimensions (e.g., stress), while the horizontal axis denotes time. These transformed dimensions of experience into time series can be then analysed in parity with neurocognitive measurements and combination with daytime and sleep-related activity, physiological parameters, as well as with individual’s brain activity, measured via portable EEG headbands. For the purpose of the current study, we collaborated with autistic stakeholders in order to identify meaningful TET dimensions (i.e., 12 dimensions; e.g., attention, boredom, social anxiety, rumination).

The main research question of this proof-of-concept study is whether an association can be determined between individual participants’ patterns of sleep and circadian behaviour, sensory reactivity and experience of stress. In a personalised medicine perspective, we aim to build on an individualised approach to collect rich data sets from individuals over a period of time in an ecological setting (i.e., fully home-based assessment). In order to relate relevant and meaningful aspects of stress and experiences by autistic individuals in their everyday life we will combine subjective experience-based data collected on a custom-built study interface on a smartphone with objective neurocognitive and physiological data measured with two portable devices, an EEG headband for measuring brain activity during night sleep and a wristband for capturing physiological parameters of sympathetic and parasympathetic activity. The data collected with this new approach should set the foundation for the development of individualised data-driven prediction models of distress and sensory reactivity for autistic individuals in the long-run.

## Materials and methods

### Participants

We are aiming to recruit at least 40 adults (20 CG/20 ASD) (31) for analyses at the group and individual level over the next 24 months. For the single participant analyses we will perform descriptive statistics and in a subsequent phase (in the next paper) a specific series of analyses between the TET and the neural dynamics, like we have done in a previous study (36). This first level statistics will define the exact statistical models to be used in Hypotheses 1-4, but assuming minimal loss of data we have 14 instances per participant which provides a minimal of 280 datapoint per cell in the mixed models we are planning to use, considered enough for robust testing of most internal contrasts.

Inclusion criteria for both groups of participants will comprise the following: age between 18 and 65 years, normal or corrected-to-normal vision, and IQ>70 in the *revised Culture Fair Intelligence Tes*t *2* (CFT-20R) (37). Autistic individuals should have received a medical diagnosis of autism with the code 6A02.0 (ICD-11), F84.5 (ICD-10), or F84.0 (ICD-10). Further psychiatric comorbidities (ICD-10 F codes) in the ASD group are no exclusion criteria in general but will be recorded. No diagnosed psychiatric (ICD-10 F codes) or acute neurological disorder should be present in the control group (CG). Medication will be recorded. Autistic individuals will be recruited in Germany and the UK through the outpatient department for autism spectrum disorders at the LMU Hospital in Munich and the Cambridge Autism Research Centre (CARD), as well as through autism-related network partners.

Prior to initiating data sampling, participants will receive comprehensive study information and the consent form. Inclusion criteria will be assessed verbally and by examining demographics at this stage. Additionally, participants will undergo the *CFT-20R* test (37). If any of the defined exclusion criteria are met, participants will be ineligible to participate in the study.

### Design and procedure

The study PRE.2021.09123-0220, named as ‘Circadian and homeostatic effects on stress and daytime neurophenomenology in autism spectrum conditions’, was approved by the Cambridge Psychology Research Ethics Committee on the 15^th^ of March 2022 (see Approval Letter 1).

Additionally, an updated German version of the study, named as ‘STREAM: Stress in Autism’, was approved by the LMU Ethics Committee on the 11^th^ of May 2023 (23–0220) (see Approval Letter 2). Honouring the relevance of a participatory approach to ASD research, the study protocol was developed iteratively incorporating feedback on feasibility, intelligibility, and relevance from 6 volunteer pilot participants with and without ASD. Adjustments and advancements to the design, instructions, schedule of testing and content (e.g., dimensions tested with TET) were made during this participatory process. Participants’ written consent and demographic data will be collected on paper before commencing the study proper. We will provide full equipment, including two wearables and a compatible smartphone for the time of participation. Participants will then undergo training in the use of the equipment either online or in person. To allow for remote data collection of subjective data and to avoid collecting data on paper, a dedicated study interface was developed by members of the laboratory using *Unity 2020.3.38f.* Both German and English versions of the study interface are available for an Android operating system.

At the beginning of the study, all participants fill out the following symptom questionnaires within the study interface for sample description (Table 1): *Beck Depression Inventory* (BDI-II) (38), the short version of the *Autism Spectrum Quotient* (AQ-10) (39), the short version of the *Sensory Perception Quotient* (SPQ-10) (40), the *Pittsburgh Sleep Quality Index* (PSQI) (41), the *State-Trait Anxiety Inventory* with focus on items of trait anxiety (STAI (Trait)) (42), the *Perceived Stress Questionnaire* (PSQ) (43). Fig 1 gives a schematic overview of the study setup.

**Table 1.**
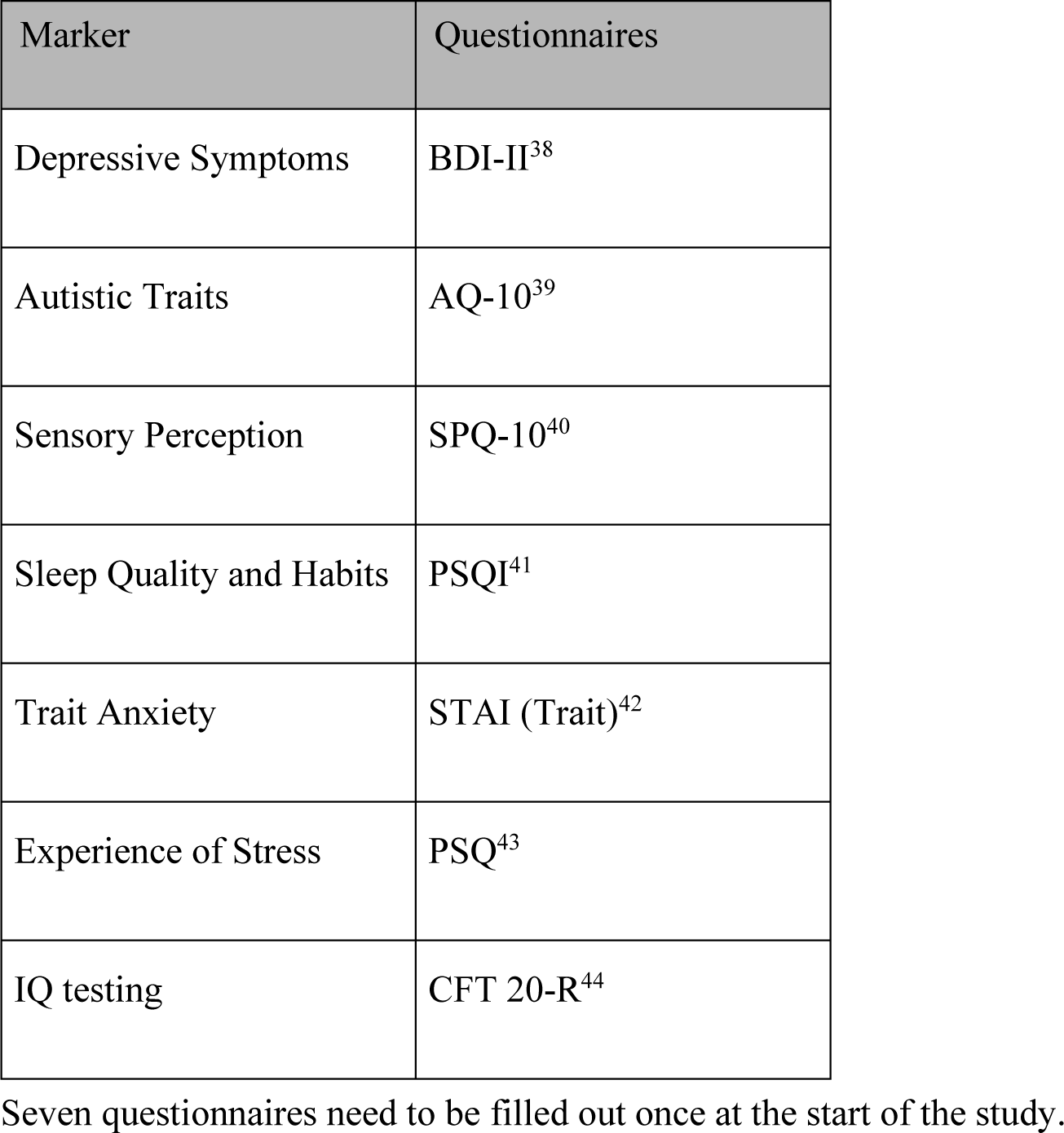
Standard questionnaires.

**Fig 1.**
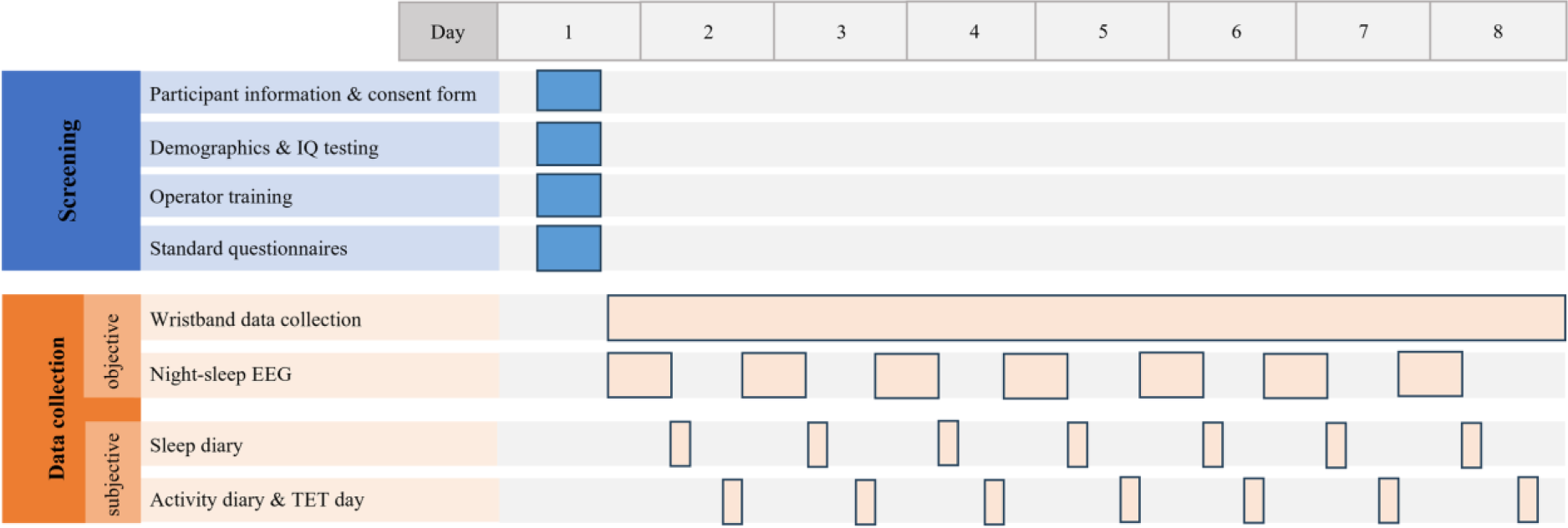
Schematic overview of the study. *Note.* The figure gives an overview of the study setup. First baseline assessments are performed. If the patient meets all inclusion criteria, is willing to participate and demonstrates proficiency in suing the devices, data acquisition starts. The timeseries shows that both objective and subjective data are measured over several days. Participants wear a wristband continuously to measure physiological data, and each night they record their sleep using an EEG-headband. In the morning, participants complete a sleep diary. In the evening, they fill out an activity diary and use the temporal experiences tracing (TET) tool to report about experiences made during the day, both by using the study interface.

#### Sleep measures

Brain activity during night sleep is being recorded using portable EEG-headbands for at least 7 nights in a row. These headbands cover frontal and occipital regions of the brain with electrodes. Signals are sampled at 250 Hz. Several neural markers will be extracted from participants’ EEG data recorded while sleeping (see Table 2). While EEG data provide objective parameters for assessing participants’ sleep quality, a sleep diary within the study interface will collect participant’s experienced sleep quality. Participants will be asked to report every morning on their night’s sleep quality.

**Table 2.** Sleep: neural parameters.

| Abbreviation | Marker | Unit |
| --- | --- | --- |
| **TST** | Total sleep time | Minutes |
| **SOL** | Sleep onset latency | Minutes |
| **MAI** | Micro arousal index | Number |
| **WASO** | Wake after sleep onset (WASO) | Minutes |
| **N1** | Stage N1 duration | Minutes |
| **N2** | Stage N2 duration | Minutes |
| **N3** | Stage N3 duration | Minutes |
| **REM** | Stage REM duration | Minutes |
| **NREM** | Stage NREM duration | Minutes |
| **N1_percentage** | Proportion of TST in stage N1 | Ratio (%) |
| **N2_percentage** | Proportion of TST in stage N2 | Ratio (%) |
| **N3_percentage** | Proportion of TST in stage N3 | Ratio (%) |
| **REM_percentage** | Proportion of TST in stage REM | Ratio (%) |
| **NREM_percentage** | Proportion of TST in stage NREM | Ratio (%) |
| **AW** | Awakenings | Number |
| **TIB** | Total time in bed | Minutes |
| **SE** | Sleep Efficiency = TST/TIB | Ratio (%) |

Neural parameters which are suggested to be extracted from participants' night sleep EEG data.

#### Physiological measures

Measuring continuously physiological parameters, such as heart rate (HR), heart rate variability (HRV), and skin conductance (SC), helps in objectively quantifying participants’ activity level of their autonomic nervous system, encompassing both sympathetic and parasympathetic activity.

HR and SC are measured using a wristband equipped with electrodes and LED lights. The wristband samples HR data at 64 Hz, while SC is sampled at 4 Hz. Participants wear the wristband throughout the day at the non-dominant hand for the time period of 7 days. Several physiological markers will be extracted as objective proxies for stress and physical activation level from participants’ wristband data (see Table 3-4). Within the study interface participants will report daily on their physical activities using a provided activity diary to give the research team some more information about the structure and environmental factors of each single day.

**Table 3.** Physiological measures: heart rate and heart rate variability.

| **1. Heart rate measures** |  |  |
| --- | --- | --- |
| Parameter | Description | Unit |
| **HR** | Mean heart rate | Beats per minute (bpm) |
| **HR_Range** | Difference between the highest and lowest heart rate | Beats per minute (bpm) |
| **SNS** | Sympathetic activity index | Index |
| **PNS** | Parasympathetic activity index | Index |
| **Stress** | Stress index | Index |
| **2. Heart rate variability (HRV): time-domain measures** |  |  |
| Parameter | Description | Unit |
| **SDNN** | Standard deviation of NN intervals | Milliseconds (ms) |
| **SDNN_Index** | Standard deviation of the average NN intervals for each 5 min segment | Milliseconds (ms) |
| **pNN50** | Percentage of successive RR intervals that differ more than 50 ms | Percentage (%) |
| **3. Heart rate variability (HRV): frequency-domain measures** |  |  |
| Parameter | Description | Unit |
| **LF_power** | Relative power of the low-frequency band (0.04-0.15 Hz) | Percentage (%) |
| **HF_power** | Relative power of the high-frequency band (0.15-0.4 Hz) | Percentage (%) |
| **LF/HF** | Ratio of low-frequency to high-frequency power | Ratio (%) |

Several heart rate and heart rate variability (HRV) measures get extracted from the blood volume pressure signal of a wristband.

**Table 4.** Physiological measures: skin conductance.

| **1. Skin Conductance (SC)** |  |  |
| --- | --- | --- |
| Parameter | Description | Unit |
| **SC_Mean** | Mean value of skin conductance | Micro siemens (uS) |
| **SC_Range** | Difference between the highest and lowest point in SC | Micro siemens (uS) |
| **SC_AUC** | Area under curve (AUC) for the SC signal | Index |
| **2. SC: Phasic Activity** |  |  |
| Parameter | Description | Unit |
| **Phasic_Mean** | Mean value of the phasic activity signal | Micro siemens (uS) |
| **Phasic_Std** | Standard deviation of the phasic activity signal | Micro siemens (uS) |
| **Phasic_Range** | Difference between the highest and lowest point in phasic activity | Micro siemens (uS) |
| **Phasic_Peak** | Number of peaks in phasic activity | Index |
| **Peak_Amplitude** | Average of peaks' amplitude | Micro siemens (uS) |
| **Phasic_AUC** | Area under curve for the phasic activity signal | Index |
| **Phasic_AUC_Percentage** | Proportion of Phasic_AUC in SC_AUC | Ratio (%) |
| **3. SC: Tonic Activity** |  |  |
| Parameter | Description | Unit |
| **Tonic_Mean** | Mean value of tonic activity | Micro siemens (uS) |
| **Tonic_Range** | Difference between the highest and lowest point in tonic activity | Micro siemens (uS) |
| **Tonic_AUC** | Area under curve for the tonic activity signal | Index |
| **Tonic_AUC_Percentage** | Proportion of Tonic_AUC in SC_AUC | Ratio (%) |
| **Phasic_Tonic_AUC** | Ratio of Phasic_AUC and Tonic_AUC | Ratio (%) |

Measured levels of skin conductance (SC) gets decomposed into phasic and tonic activity. For all three variables several parameters will be calculated.

#### Experience based measures

To ensure that the dimensions participants use to represent their daily experiences align with our research objectives, the experience dimensions were developed in collaboration with autistic advisors, leading to the following nine dimensions (see Table 5): wakefulness, boredom, sensory avoidance, social avoidance, physical tension, scenario anxiety, rumination, stress, and pain. A short description of all dimensions is given in the study interface and participants also have the option to add a personalised dimension. For a duration of 7 days, participants will fill out daily diaries within the study interface, reporting on their night’s sleep quality and activities throughout each day. Additionally, participants are requested to provide daily reports on the dynamics of specific dimensions of experiences made throughout the day by using the integrated TET tool (see Figure 2).

**Table 5.** Temporal Experience Tracing (TET): overview and description of daily dimensions.

| Number | Experience | Question (range) | Description |
| --- | --- | --- | --- |
| 1 | Wakefulness | How alert did you feel during the day?<br>(not alert – alert) | High levels of wakefulness are synonymous with the feeling of alertness and physiological arousal. Low wakefulness can mean you are feeling quite drowsy or even falling asleep entirely (e.g., taking a nap during the day). |
| 2 | Boredom | How bored did you feel during the day?<br>(not bored – bored) | Boredom is an adaptive response to situations, where little novel information is available, there is little progress or change, or there is a lack of general mental and physical activity, while a desire for such activity is present. It can be an unpleasant affective reaction in face of surroundings that do not offer anything new or interesting that will allow us to learn about or interact with the world. |
| 3 | Sensory avoidance | Were you avoiding stimulation of your senses (touch, taste, sound etc)?<br>(not avoiding – avoiding) | Sensory avoidance is referring to the degree you were trying to avoid specific sensory stimuli during the day. This dimension refers to all possible sensory qualities (e.g. sight, smell, hearing, taste and touch). It is also possible that your avoidance of sensory input is switching<br>between specific qualities over a single day. It might happen for instance, that you are avoiding visual input while you are in a new environment, but when being at home you focus on the hearing sense or on the taste when having dinner. |
| 4 | Social avoidance | Were you avoiding social interactions (virtual and/or in person)? (not avoiding – avoiding) | Social avoidance is referring to the degree you were trying to avoid social contact and/or social interaction during the day. Examples might include: you were staying at home, cancelled a meeting, didn't text someone back or you were avoiding phone calls etc. |
| 5 | Physical tension | How physically tense did you feel throughout the day?<br>(not tense – tense) | This item describes the intensity to which specific body parts or your whole body were feeling physically tense during the day. The feeling of physical tension can increase or decrease over the day, but it can also change the part of the body where you refer this tension to (e.g. muscles, neck, head). |
| 6 | Scenario anxiety | How much were you lost in thoughts worrying about present and future events (e.g. scenario planning,<br>trying to solve problems)?<br>(not worrying – worrying) | Scenario anxiety is asking you to evaluate how much you were lost in thoughts worrying about present and future events. This can mean scenario planning, as well as trying to solve problems. |
| 7 | Rumination | How much were you lost in thoughts worrying about past events (e.g., rumination, replaying interactions)?<br>(not worrying – worrying) | This item is asking how much you were worrying and thinking about the past during the day. |
| 8 | Stress | How stressed did you feel during the day?<br>(not stressed – stressed) | This dimension is asking you to which extent you were feeling stressed over the day. |
| 9 | Pain | How strong did you feel physical pain?<br>(none – very strong) | This dimension is asking you to which extent you were feeling physical pain during the day. |
| 10 | Personalised dimension |  | This dimension is not further specified. If you experience that a specific quality or item is not covered by the 9 items mentioned before, feel free to define this item as you wish. Please add a short description to the y-axis. |

Nine dimensions are defined for the Temporal Experience Tracing (TET) tool referring to participants' daily experiences. Participants are free to add another personalised dimension.

**Fig 2.**
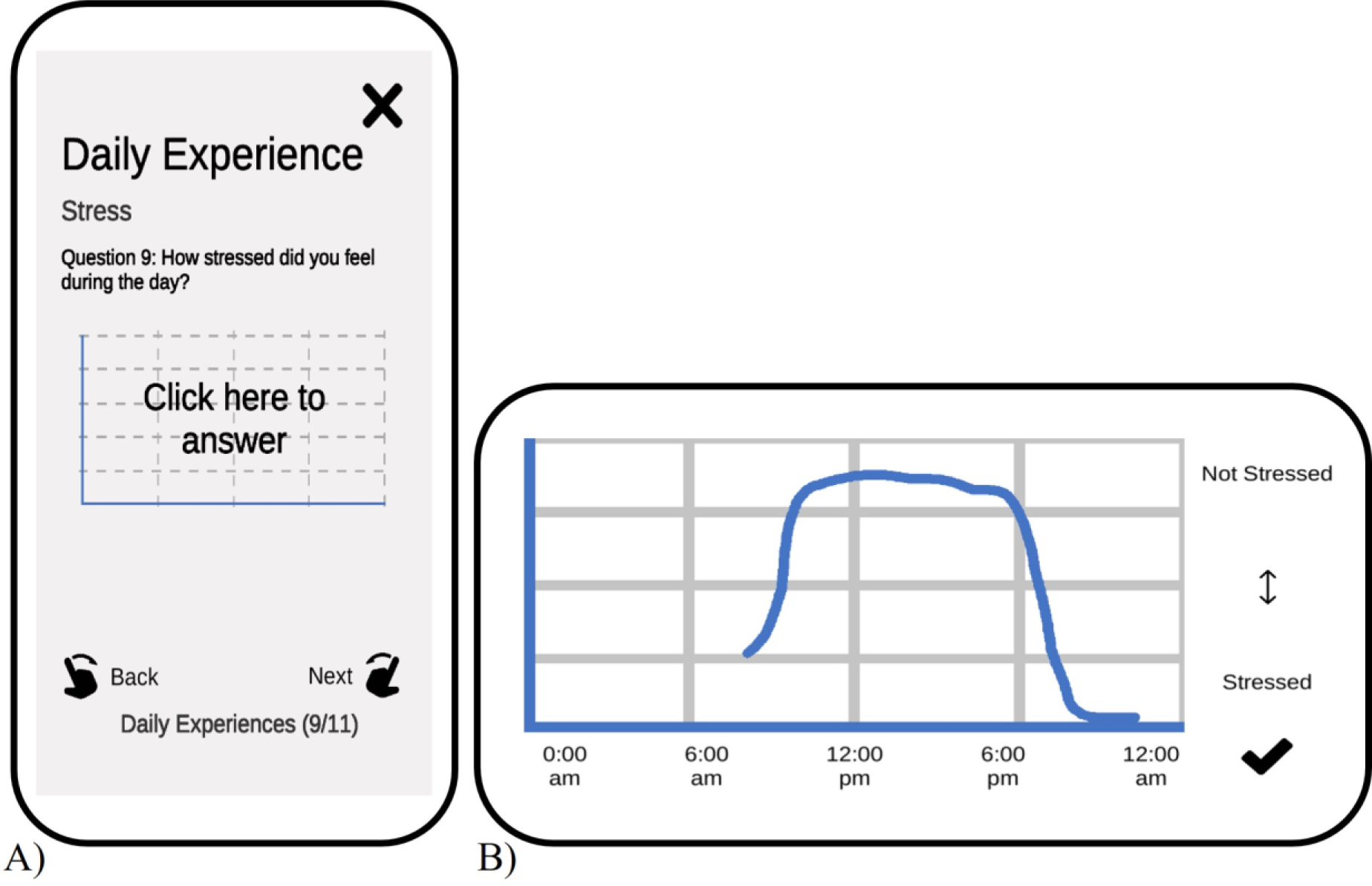
Temporal Experience Tracing – an example from the study interface. *Note.* A) Tracing the dynamics of daily experiences at the end of the day is one task in the project. In this example participants are asked to trace their feelings of ‘Stress’ throughout the day. B) Example of an answer for this question. Participants can digitally draw the graph within the study interface.

### Data management

Personally identifiable information and clinical data from participants will be kept separately from the experimental data and will not be provided to individuals beyond the research team. To access the study interface on the study smartphone and enter responses, participants are assigned a randomly generated numerical identifier. All answers of the study interface are stored in a secure university data server using the numerical identifier.

The code for decrypting the personal data is only available to the database administrator and is locked on a local computer that is not connected to the network. The data management complies with the requirements of the Declarations of Helsinki (2013) (44) and the General Data Protection Regulation (GDPR) (45).

For both portable devices, the EEG headband and the wristband, the manufacturers provide a dedicated smartphone application to transfer data from the device via Bluetooth connection to a secure, password-protected cloud system. Both apps are already installed on the study smartphone when given to the participant. Each participant will receive an individual, pseudonymized numerical code to log into those two smartphone applications. Access to the cloud systems is restricted to members of the research team, allowing them to download and securely store the data on a secure University server.

### Risk management

Participants will receive participant information and consent form before starting the study, which include safety instructions for the devices from the manufactures’ instruction sheets, among other details. Additionally, all participants will receive operator training on device usage and have access to an instructional video included in the study interface, available for viewing at their convenience. Participants are encouraged to record data continuously over several days to understand potential effects of a cumulative sleep-deficit on daytime sensory processing and subjective experiences of stress. All data is collected non-invasively, and there are no actual associated risks for participants. Participants will be informed that they can stop data collection and study participation at any time without providing a reason. However, they can also interrupt the study protocol for a short break (e.g., due to acute illness). The study protocol of 7 days can be extended if data collection is unsuccessful in more than two days and the participant is willing to extend data acquisition. Data collection will be stopped if participants are unable to start data collection within one week of receiving the devices or if the break between data collection days exceeds one week. Participants can contact the research team via email or phone for any questions or technical issues with the devices. Spare devices will be available, and replacements will be sent if needed.

The experimenter will monitor data quality and if it is unsatisfactory, the researcher will guide the participant to mitigate any issues. Nested statistical models, which are robust regarding missing data, will be used for analysis.

Some participants may lack the digital skills needed for the study. Digital skills and task comprehension will be discussed at the start of the experiment. We only include participants who demonstrate sufficient digital and wearables usage skills to avoid too high burden for those without digital literacy skills. Participants will receive regular support from the experimenters throughout the study through fixed virtual meetings.

### Data analysis plan

#### Reported sleep quality

Given the high prevalence of sleep disorders (46) in autism, we assume significant differences in subjective ratings and measures of sleep quality when analysing participants’ sleep diaries (29).

H1.1) We expect autistic individuals to report longer sleep onset latency, more night awakenings, lower sleep duration and lower sleep efficiency in the sleep diary compared to individuals of the control group.

H1.2) **Objective sleep quality**. We expect differences between autistic individuals and control individuals in their EEG sleep patterns (REM and N3 time, awakening, sleep onset latency and total sleep) (29–30).

H2.1) **Peripheral stress measures**. We hypothesise that autistic individuals show higher sympathetic activity and stress indices throughout the day, lower heart rate variability and more phasic activity in their SC time series compared to the CG.

H2.2) **Subjective stress measures.** We expect autistic individuals to report higher levels of subjectively experienced daytime stress compared to the CG.

H3) **ASD subjective measures**. We predict that autistic individuals will show higher absolute values and a higher variance in the graphs of the TET tool for daily dimensions when reporting about dimensions that are linked to autism specific behaviour (i.e., social avoidance, sensory avoidance).

H4) **Daily subjective stress association with sleep quality**. We expect the TET parameters of stress to be associated with the following night’s self-reported and objectively measured sleep quality.

Additional exploratory hypotheses might be tested and preregistered in the project’s OSF page (https://osf.io/e9qtu/).

For all analyses, the daily TET data corresponding to each dimension (e.g., stress, sensory hypersensitivity) will be sampled every 20 minutes, resulting in approximately 40 data points per day. Between-participant and within-participant variations are of interest. Significance for statistical tests is defined at p<.05. Results will be corrected for multiple comparison using FDR.

We will test H1.1 and H1.2 using both data from the sleep diary and EEG recordings, objective and subjective sleep quality. The neural data will be analysed to determine the hypnogram and infer sleep depth and quality. Control and autistic groups will be compared using mixed level modelling at the group analyses levels both with frequentist stats and bayes information criterion.

For the peripheral stress measures (wristband) (H2.1) we will also perform mixed level modelling between autistic individuals and control group expecting higher sympathetic activity and stress indices throughout the day in the objective measures.

For H2.2 and 3, subjective daily measures of stress and ASD related dimensions of experience, we will use coefficient of variation and trend analyses on the TET to obtain a summary measure of the trace that better represent the dynamics than the average. Using those we will compare between groups all measures together in a mixed model, also using group, single participant, the measures and their covariations as factors in the model.

To test the association between daily subjective stress and sleep quality (H4), we will test the statistical dependencies between the daily TET of stress and the objective and subjective sleep quality measures the following night using mixed regressions between and within groups.

## Discussion

This study addresses the relationship of several common co-occurring conditions in ASD: sleep disorders, sensory reactivity, and stress. The study protocol described herein includes several aspects of novelty in the field of autism research concerning the design and research methods. First, subjective experiences are quantified on an individual level and related to psychophysiological and neurophysiological (i.e., objective) measurements. Second, data are collected over a prolonged period of time allowing for analysis of individual patterns of interrelation between circadian activities and sleep, sensory reactivity, and stress. Third, the relevance of experience dimensions were discussed with and based upon feedback from autistic individuals, including an autistic co-author of this protocol. In addition, participants will be assessed during their daily lives at home using wearables and digital measurements that enable us to apply lab-level control within the ecological setting of people’s real-life experience and thereby increase relevance. Thus, we expect clinical relevance of the study approach and to learn more about the individual aspects of stress experience in ASD.

Aside from the methodological advantages and the importance of the addressed research questions in the field of autism, the study design has following limitations. First, a personalised approach over a longer period implies that it is more difficult to collect data from a large sample size. It is crucial to acknowledge that sampling bias may arise, particularly concerning sensory processing, which is one of the core aspect of the current study. The authors recognize that this bias results from the inclusion only of those who are capable and willing to wear the equipment. Nevertheless, we will learn important and new aspects about the interrelation of individual stress experience, sensory reactivity as well as circadian and sleep irregularities by focusing on personalised patterns gained from rich individual longitudinal data sets (47). Future studies could, on the basis of the results of the current study, apply a more tailored individual approach making similar studies lighter and more accessible to a wider spectrum of individuals.

Second, we aim to investigate the potential of objective and subjective outcome variables for classification of clusters within and between individuals with and without ASD. Questions about the directionality of effects and etiological pathways remain a target for future prospective studies and are outside the scope of the current study design. Nevertheless, the current study design will enable hypotheses of causal relationships to be generated and specifically tackled in future tailored studies. Given the heterogeneity in the autism phenotype and with respect to constantly changing environmental factors, beside from individual, time, and context dependent factors (e.g., unknown coping strategies, comorbidities, social support), we might not derive a general pattern across all participants. Still, we expect to be able to assess individual interrelationships that can inform about factors relevant for prediction of stress and sensory reactivity. In this respect, we account for several potential cofactors in our study by using different diaries and addressing several dimensions of experiences, besides giving participants extra space to report any irregularity of the day and their state.

Third, given TET is a method in which participants graph intensity of their experiences retrospectively, it might lead to memory biases. Recall biases are not specific for autistic individuals and could likewise occur in control participants and this issue similarly concerns traditional alternatives such as rating scales. As a precaution, we advise participants to draw their experiences in a timely fashion to reduce the recall bias in this task. We are convinced though that TET offers a great opportunity to get more insight into the dynamics of subjective experiences, something that cannot be derived from standard rating scales (48).

Fourth, portable devices are used for data acquisition in this research project and we are aware that data quality might be lower due to motion artefacts and lower sampling frequencies compared to lab-controlled assessments, where devices such as an electrocardiogram (ECG) for patients’ HR or polysomnography (PSG) for assessing sleep quality are used. Additionally, the EEG headband has a scalp coverage limited to a few frontal and occipital electrodes, which yields EEG data with low spatial resolution. Despite of these limitations, the portable devices used in the current study allow for capturing naturalistic behaviour, experiences, and physiological responses of individuals in their daily and relevant environment. Wearables offer this possibility and flexibility. We address upcoming issues in data quality by giving participants regular feedback about the quality of their recordings and discuss individual adjustments in the process of data acquisition, in addition to adapted pre-processing methods when analysing raw data.

## Conclusions

The current study protocol will deliver insights into individual patterns of objective and subjective stress experienced by individuals with ASD and relate it to their patterns of circadian activity and sleep as well as sensory reactivity. The results will help tailor future interventions to tackle stress prediction for better mental health outcomes in the autistic population.

## Data Availability

As the paper presents a study protocol, no data is yet available for sharing.

## Acknowledgements

We express our gratitude to the autistic and non-autistic pilot-participants whose feedback, insights and collaboration played a crucial role in refining and advancing the study protocol.

